# Anaemia among mother-child dyads in India: trends, drivers, and future projections

**DOI:** 10.1101/2025.01.23.25321031

**Authors:** Sarang Pedgaonker, Trupti Meher, Monali Gupta, Suman Chakrabarti, Phuong Hong Nguyen, Shri Kant Singh, Rasmi Avula, Laxmi Kant Dwivedi, Aditi, Samuel Scott

## Abstract

Anaemia among mothers and their children is a widespread public health challenge with profound consequences for individuals and societies. While anaemia has been studied separately in women and children, there remains a literature gap examining anaemia in mother-child dyads, limiting insights on interventions that may simultaneously address anaemia in both groups. Our study examines trends and drivers of anaemia among mother-child dyads (N=408,342) in India using nationally-representative data from 2006 to 2021 and estimates the potential future reduction in anaemia based on changes in selected drivers. We employed descriptive statistics, multivariable logistic regression and population attributable fraction (PAF) analysis. The co-occurrence of anaemia among mothers-child dyads decreased from 35% in 2006 to 33% in 2016, but increased to 37% in 2021. Subnational analyses revealed varying trends by states, with Delhi showing the highest increase (17% to 32%) and Sikkim the largest decrease (29% to 16%) between 2006 and 2021. Maternal education, regular consumption of non-vegetarian food and green leafy vegetables, consumption of iron folic acid supplements, utilization of government health services, and improved sanitation at both household and community levels were associated with lower likelihood of anaemia among mother-child dyads. The cumulative PAF suggested that addressing these factors collectively could reduce anaemia prevalence among mother-child dyads by 18% to 28% (under different scenarios) by 2030. The study underscores the need for comprehensive, multi-sectoral interventions targeting both maternal and child health to effectively combat anaemia in mother-child dyads.

**Key messages:**

- The co-occurrence of anaemia among mother-child dyads in India increased from 33% in 2016 to 37% in 2021. More effective strategies and interventions to combat anaemia are needed.
- Realistic improvements in maternal education, dietary practices, toilet facilities and reduction in community open defecation could reduce anaemia prevalence among mother-child dyads by 18% by 2030.
- More optimistic scenarios could reduce anaemia burden by 28% by 2030, with improvements in education and sanitation being critical.

## Introduction

Anaemia is a public health concern linked to poor pregnancy outcomes, chronic fatigue, stunted growth, poor cognitive and motor development in children, reduced work productivity, and increased risk of morbidity and mortality (Murray-Kolb et al., 2009; WHO, 2023). Anaemia is not a disease in itself but a manifestation of various underlying causes, including nutritional deficiencies, chronic diseases, genetic disorders and infection (WHO, 2023). Its impact extends beyond individuals, with social and economic consequences (Alcázar, 2013).

Globally, anaemia affects approximately 1.92 billion people or one-fourth of the world population (GBD 2021 Anaemia Collaborators, 2023), with women of reproductive age and under-five children being the most affected (Stevens et al., 2013; Shekar et al., 2017). It is estimated that 40% of all children aged 6–59 months and 30% of women aged 15–49 years are anaemic (WHO, 2023) and South Asia and sub-Saharan Africa bear the highest burden (Biradar, 2023). Poverty, malnutrition, high rates of infectious diseases, and limited access to healthcare services (IFPRI, 2015; Black et al., 2013; Agrawal et al., 2023) contribute to the high burden of anaemia in these regions (WHO, 2023).

In India, anaemia among mothers and children has fluctuated in past decades (Singh et al., 2023), with little progress overall despite various governmental and non-governmental efforts to reduce the problem. According to India’s 2019-2021 National Family Health Survey (NFHS-5), 57% of women of reproductive age and 67% of under-five children are anaemic (IIPS, 2021). The persistent high prevalence of anaemia in India suggest that existing interventions may be insufficient or inadequately implemented, necessitating a rethink on how to effectively address anaemia in this population.

While anaemia among women and children has been studied separately, there remains a literature gap examining anaemia in mother-child dyads. Given that mothers and children share dietary habits and living conditions, anaemia in mother-child pairs is likely driven by a common set of factors. For instance, food intake in children is highly dependent on food intake by their mother (Kueppers et al, 2018) and children of anaemic mothers are more likely to be anaemic and face developmental challenges than children of mothers without anaemia (Heesemann et al., 2021; Berner et al., 2014). Studying anaemia in mother-child dyads may yield insights that help break the intergenerational transmission of anaemia.

Previous research has shown that anaemia among mother-child pairs is more prevalent in poorer households in India, highlighting the need for targeted interventions in impoverished communities Biradar (2023). A study in Bangladesh also emphasized the importance of considering both children and mothers in anaemia-related health programs as well as cohort-specific tailored interventions to effectively reduce the anaemia burden (Khan et al. (2020).

Building on this evidence, our study aimed to investigate the trends and variations in anaemia prevalence among mother-child dyads in India over the past decade and a half. By analyzing data from multiple time periods and subnational regions, we sought to provide a comprehensive understanding of the factors influencing anaemia and to inform the development of more effective public health strategies. The objectives of the present study were to 1) examine the co-occurrence of anaemia among mother-child dyads in India from 2006 to 2021, 2) identify key drivers of anaemia in this population, and 3) estimate future reductions in anaemia in this population under different scenarios of changes in drivers of anaemia. We hope that the findings from this study can provide a roadmap for long-term efforts to reduce anaemia.

## Methods

### Data source

Data from three rounds of India’s National Family Health Survey (NFHS) were used: NFHS-5 (2019-2021) (IIPS & ICF, 2021), NFHS-4 (2015-16) (IIPS & ICF, 2017), and NFHS-3 (2005-06) (IIPS, 2007). These surveys were conducted by the International Institute for Population Sciences (IIPS), under the stewardship of the Ministry of Health and Family Welfare (MoHFW), Government of India. NFHS contains comprehensive data on population, health, nutrition, family planning services, women’s autonomy, domestic violence and a range of underlying determinants. NFHS is India’s equivalent to Demographic and Health Surveys conducted in other countries.

NFHS adopted a stratified two-stage sampling design. The first stage involved the selection of primary sampling units (PSUs) – villages in rural areas and census enumeration blocks in urban areas – using population proportion to size sampling. The second stage involved the random selection of 22 households from each PSU using equal probability systematic sampling, and a complete household mapping and listing operation was carried out prior to the main survey. Detailed survey sampling procedures and questions are available in the final reports of NFHS-3 (IIPS, 2007), 4 (IIPS & ICF, 2017) and 5 (IIPS & ICF, 2021). Since this study focuses on anaemia among mother-child dyads, the analysis was confined to women aged 15-49 years and their children aged 6-59 months, a total sample size of 28,983, 195,993 and 183,366 in NFHS-3, 4 and 5, respectively.

### Outcome variable

The outcome of interest was anaemia among mother child dyads, assessed using haemoglobin (Hb) concentrations from capillary blood samples. A portable HemoCue Hb 201^+^ analyzer was used by health investigators to measure Hb concentrations, obtained by finger pricking for adults and heel pricking for children aged 6-11 months. Hb level was adjusted for smoking (in women) and altitude (in women and children) in enumeration areas above 1000 meters. The level of anaemia among mother and children were defined according to WHO guidelines (WHO, 2024) as follows:

|  | Hb level |  |  |  |
| --- | --- | --- | --- | --- |
|  | Any anaemia | Severe anaemia | Moderate anaemia | Mild anaemia |
| Children (6-23 months) | < 10.5 g/dl | < 7.0 g/dl | 7.0 – 9.4 g/dl | 9.5 – 10.4 g/dl |
| Children (24-59 months) | < 11.0 g/dl | < 7.0 g/dl | 7.0 – 9.9 g/dl | 10.0 – 10.9 g/dl |
| Mother | <12.0 g/dl | < 8.0 g/dl | 8.0 – 10.9 g/dl | 11.0 – 11.9 g/dl |

### Explanatory variables

We explored drivers of anaemia across maternal, child and household levels. Maternal factors included age, education (primary: schooling 1-5 years; secondary: schooling 6-12 years; higher: schooling >12 years), mass media exposure, food consumption (consumption of animal food, and consumption of green leafy vegetables), antenatal care (ANC) visits, preceding birth interval, postpartum haemorrhage, iron folic acid (IFA) consumption (100+ days) and deworming. Mass media exposure encompasses the frequency of reading newspapers/magazines, listening to the radio, and watching TV. Responses of ‘almost every day’ and ‘at least once a week’ were coded as 1, while ‘not at all’ and ‘less than once a week’ were coded as 0. The final variable was created by summing the frequency of exposure to these three media types, categorizing them as ‘no’ for zero exposure, ‘low’ for exposure to one medium, and ‘high’ for exposure to more than one medium. Maternal food consumption was measured by asking women how often they consume the food items (never/occasionally, weekly, daily). Child-level factors included age, sex, siblings under five years of age, low birth weight/size, receipt of food supplements (take home rations) through the Integrated Child Delivery Service (ICDS) scheme, and morbidity. Child morbidity included fever, diarrhoea and acute respiratory infection (ARI), which were assessed based on maternal recall of symptoms in the two weeks prior to the survey. This indicator was a dummy for having at least one of the three conditions. Household size, place of residence, caste, asset index, drinking water facility, toilet facility, community open defecation and cooking fuel were included under household-level factors. Community open defecation was measured as the average proportion of households in a primary sampling unit that reported practising open defecation (Hamlet et al., 2023).

### Data analysis

To examine trends in the co-occurrence of anaemia, mother-child dyads were initially divided as follows: both non-anaemic, mother anaemic & child non-anaemic, mother non-anaemic & child anaemic, and both anaemic. These dyads were then categorized into four groups based on increasing severity of anaemia: both non-anaemic (category 1), one or both members with mild anaemia (category 2); one member non-anaemic and the other member moderately or severely anaemic (category 3); both members anaemic, with at least one member moderately or severely anaemic (category 4). In addition to national trends, to visualize state-level variability in trends, maps were created for three survey rounds. The association between potential drivers (explanatory variables) and outcomes was examined using binary logistic regression for each survey round.

Population attributable fraction (PAF) analysis was used to estimate future reductions in anaemia under different scenarios of changes in drivers of anaemia. PAF is commonly defined as the proportional reduction in average disease risk over a specified time interval that would be achieved by eliminating the exposure(s) of interest from the population while the distributions of other risk factors in the population remain unchanged (Rockhill et al., 1998). The following formula was used to determine the PAF of a particular risk factor.

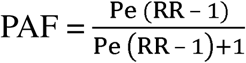

Where, P_e_ = proportion of population exposed to risk factor and RR = Risk ratio.

In the study, the adjusted odds ratio (*OR*) was used in place of *RR.* The value of the PAF depends on the prevalence of the risk factor and the strength of its association with the outcome variable.

Projected prevalence rates for each risk factor were calculated for the year 2030 under realistic and optimistic scenarios. The realistic scenario projections for 2030 were based on the average annual rate of change observed between NFHS-3 and NFHS-5. This rate reflects the typical progression of each risk factor over time. Optimistic scenarios were then derived by assuming higher annual rates of change (3% and 5% deviation from the average annual rate of change used in the realistic scenario), representing more favourable future conditions.

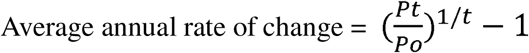

Where, P_t_ = prevalence in NFHS-5; P_o_ = prevalence in NFHS-3; t = number of years between the surveys (15 years).

The PAF for each risk factor was then estimated based on its projected prevalence, considering both individual and combined effect on anaemia among mother-child dyads.

## Results

### Trends in anaemia among mother-child dyads at national and subnational levels

There were small changes over time in the prevalence of anaemia among mother-child dyads at the national level (**Figure 1, Panel A**). In about one-third of dyads, both the mother and child were anaemic, while about one-quarter of dyads were non-anaemic for both members. In about 40% of dyads, either the mother or child was anaemic, with the other member being non-anaemic. Further breakdown into categories based on anaemia severity showed limited change over the fifteen-year period studied (**Figure 1, Panel B**). The percentage of dyads in the most severe category (both members anaemic, with at least one having moderate or severe anaemia) declined from 29% in 2006 to 26% in 2016 and then increased to 30% in 2021.

**Figure 1:**
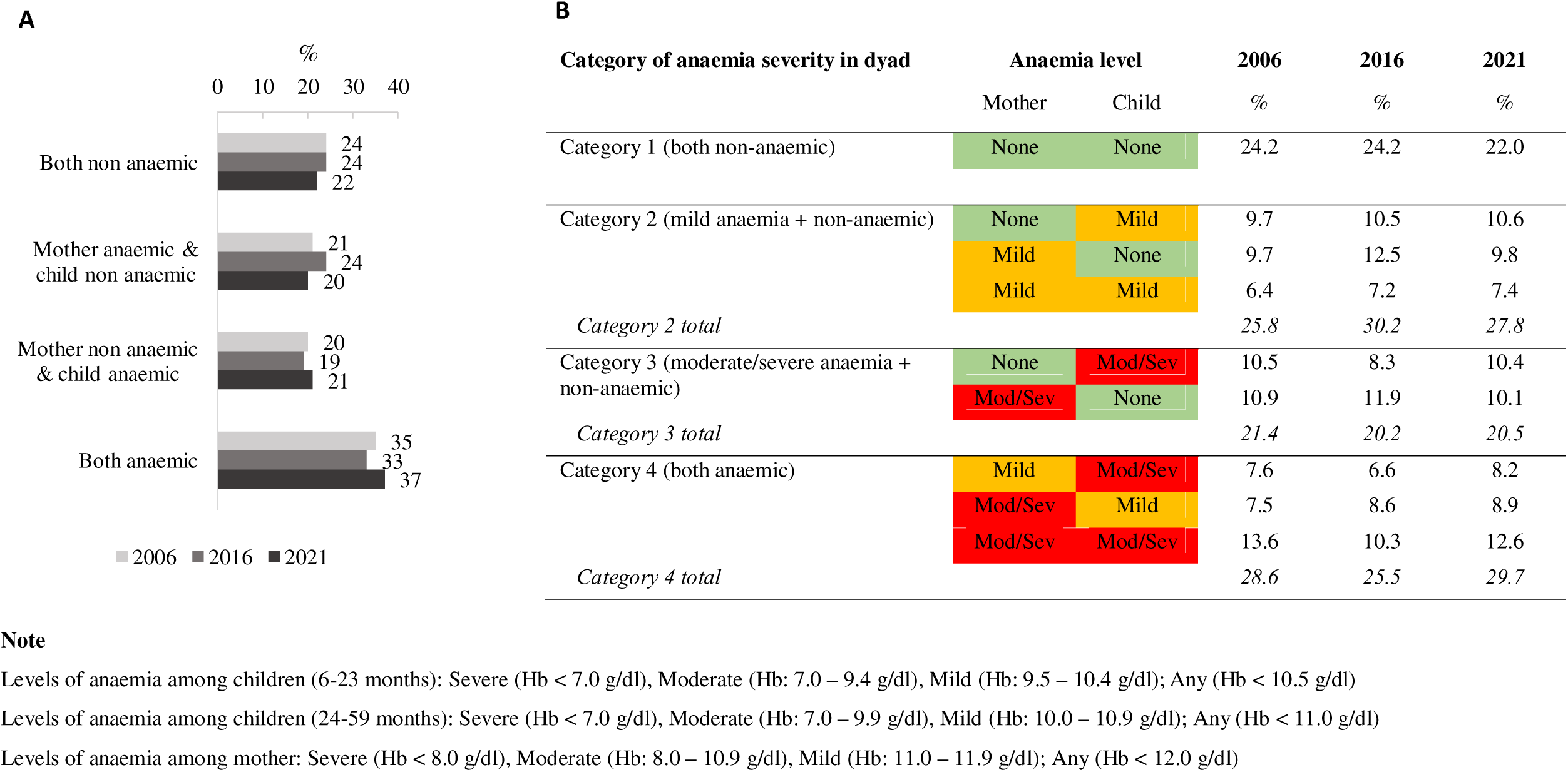
Combinations of anaemia prevalence among mother-child dyads in India, 2006, 2016, 2021

Subnational analysis revealed that the number of states and union territories (UTs) where over 40% of the dyads were classified as both anaemic decreased between 2006 and 2016, but increased between 2016 and 2021 (**Figure 2, Panel A**). Thirteen states/UTs showed a rise in the prevalence of anaemia among mother-child dyads from 2006 to 2021. Delhi reported the highest increase (17% to 32%), whereas the state of Sikkim reported the highest decline (29% to 16%) during this period. (**Table S1**). **Table S2** shows the change in the percentage of dyads for each anaemia category. From 2016 to 2021, there was a decrease in the number of districts (from 64 to 35 districts) where less than 20% of the dyads were classified as both anaemic and a substantial increase in the number of districts (from 296 to 438 districts) where over 40% of the dyads were classified as both anaemic (**Figure 2, Panel B**).

**Figure 2:**
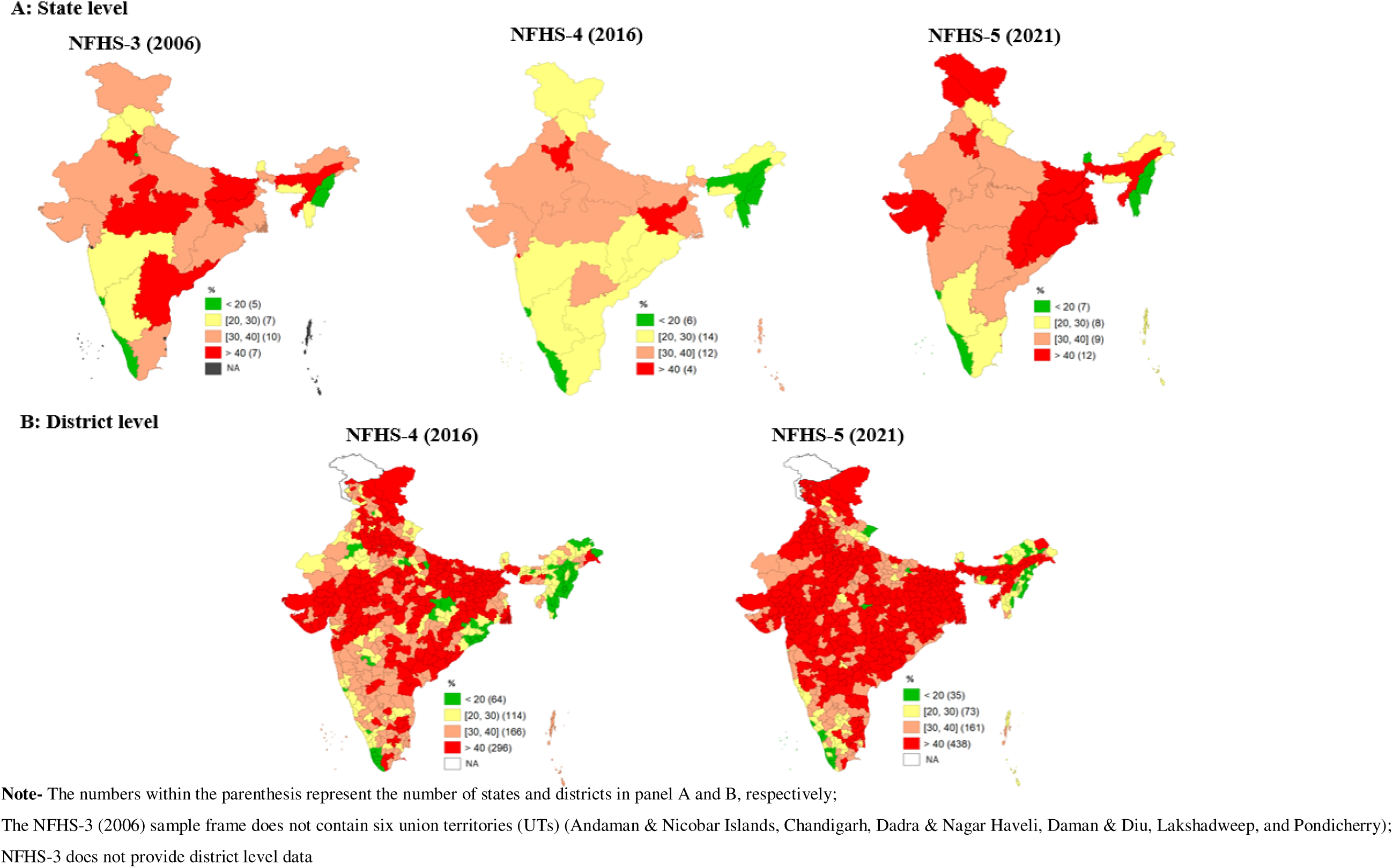
Prevalence of anaemia among mother-child dyads (both anaemic) at sub-national level, 2006, 2016, 2021

### Factors associated with anemia among mother-child dyads

From 2006 to 2021, there were increases in the percentage of mothers with higher education (8% to 16%), who consumed egg/fish/meat at least weekly (47% to 54%), consumed IFA for more than 100 days (21% to 44%) and received deworming medication during the previous pregnancy (4% to 32%) (**Table 1**). The percentage of children receiving ICDS food supplementation (using a 12-month recall period) also increased from 27% in 2006 to 66% in 2021. Nearly two-thirds of the sample lived in rural areas. There were also increases in the percentage of households with improved toilet facilities (45% to 74%) and using clean cooking fuel (27% to 51%).

**Table 1:** Maternal, child, and household characteristics, 2006, 2016, 2021.

| Background characteristics | NFHS-3 (2006)<br>(N= 28983) | NFHS-4 (2016)<br>(N= 195993) | NFHS-5 (2021)<br>(N=183366) |
| --- | --- | --- | --- |
|  | % | % | % |
| **Mother's characteristics** |  |  |  |
| Age (15-24 years) | 41 | 32 | 30 |
| Age (25-34 years) | 52 | 59 | 61 |
| Age (35-49 years) | 7 | 9 | 9 |
| Higher education (>12 years schooling) | 8 | 11 | 16 |
| Low media exposure <sup>a</sup> | 36 | 45 | 41 |
| High media exposure | 30 | 22 | 10 |
| Weekly consumption of egg/fish/meat | 47 | 50 | 54 |
| Weekly consumption of green leafy vegetables | 29 | 39 | 38 |
| Daily consumption of green leafy vegetables | 64 | 47 | 53 |
| 4+ ANC visits | 40 | 41 | 43 |
| Birth interval >23 months | 48 | 46 | 46 |
| Postpartum haemorrhage | 13 | 20 | 23 |
| 100+ days IFA consumption | 21 | 30 | 44 |
| Received Deworming | 4 | 18 | 32 |
| **Child's characteristics** |  |  |  |
| Age (6-11 months) | 14 | 12 | 12 |
| Age (12-23 months) | 22 | 22 | 21 |
| Age (24-59 months) | 64 | 66 | 67 |
| Male | 54 | 53 | 53 |
| Female | 46 | 47 | 47 |
| Siblings <5 years | 42 | 40 | 38 |
| Low birth weight/size | 21 | 17 | 17 |
| Received ICDS food supplementation | 27 | 52 | 66 |
| Morbidity in last 2 weeks <sup>b</sup> | 22 | 19 | 18 |
| **Household characteristics** |  |  |  |
| Household size has >5 members | 57 | 56 | 55 |
| Rural | 66 | 71 | 72 |
| Scheduled caste (SC)/Scheduled tribe (ST) | 29 | 32 | 33 |
| Other backward classes (OBC) | 38 | 44 | 43 |
| Asset index (poor) | 50 | 36 | 35 |
| Asset index (middle) | 27 | 28 | 33 |
| Asset index (rich) | 24 | 36 | 32 |
| Piped drinking water <sup>c</sup> | 43 | 40 | 42 |
| Improved toilet facility <sup>d</sup> | 45 | 51 | 74 |
| Community open defecation <sup>e</sup> | 48 | 43 | 22 |
| Clean cooking fuel <sup>f</sup> | 27 | 36 | 51 |

<sup>a</sup> Mass media exposure was created by summing the frequency of exposure to the three media types: reading newspapers/magazines, listening to the radio, and watching TV and then categorizing them as 'no' for zero exposure, 'low' for exposure to one medium, and 'high' for exposure to more than one medium
<sup>b</sup> Morbidity encompasses at least one of the three conditions: diarrhea, fever and acute respiratory infection (ARI)
<sup>c</sup> Piped drinking water includes piped water, public taps/standpipes
<sup>d</sup> Improved toilet facilities include any non-shared toilet of the following types: flush/pour flush toilets to piped sewer systems, septic tanks, pit latrines, or an unknown destination; ventilated improved pit (VIP)/biogas latrines; pit latrines with slabs; and twin pit/composting toilets
<sup>e</sup> Community open defecation was measured as the average proportion of households that reported practising open defecation in a primary sampling unit
<sup>f</sup> Clean cooking fuel includes electricity, LPG/natural gas, biogas
Indicators such as ANC visits, preceding birth interval, 100+ days IFA consumption, received deworming are for the last birth

In the regression analysis examining associations between determinants and the co-occurrence of anaemia in mother-child dyads for each round of data separately, dyads in which the mother had completed her higher education were less likely to be anemic in 2006 (adjusted odds ratio (AOR)=0.50, 95% confidence interval (CI): 0.34-0.73), 2016 (AOR=0.57; CI:0.53-0.62) and 2021 (AOR=0.58; CI:0.54-0.62) compared to dyads in which the mother had no education (**Table 2**). At least weekly consumption of egg/fish/meat was linked to a reduced likelihood of anaemia in 2016 (AOR=0.77; CI:0.74-0.80) and 2021 (AOR=0.91; CI:0.84-0.94). In 2016, consumption of green leafy vegetables was associated with decreased odds of anaemia (daily AOR=0.72; CI:0.69-0.76). Dyads in which mothers consumed IFA supplements for more than 100 days were less likely to be anaemic (AOR=0.92; CI:0.88-0.95) compared to dyads where the mother did not consume IFA for at least 100 days. Receipt of deworming medication was associated with higher odds of anaemia in 2016 (AOR=1.06; CI:1.01-1.11) and 2021 (AOR=1.04; CI:1.04-1.12), presumably due to program targeting. Dyads where children had siblings under the age of 5 years and where the child was low birth weight were more likely to be anaemic than their counterparts. Having an improved toilet facility at home was associated with lower odds of anaemia among mother-child dyads in 2016 (AOR=0.85; CI:0.81-0.89) and 2021 (AOR=0.91; CI:0.87-0.96). Higher community-level open defecation was associated with increased odds of anaemia (1.6 to 3 times) among both mothers and their children at all three time points.

**Table 2:** Association between selected factors and anaemia among mother-child dyads (both anaemic), 2006, 2016, 2021.

| Background characteristics | Adjusted Odds Ratio (95% CI) |  |  |
| --- | --- | --- | --- |
|  | NFHS-3 (2006) | NFHS-4 (2016) | NFHS-5 (2021) |
| **Mother's characteristics** |  |  |  |
| **Age** |  |  |  |
| 35 y & above | Ref. | Ref. | Ref. |
| 25-34 y | 1.36** (1.09-1.69) | 1.11*** (1.05-1.18) | 1.21*** (1.15-1.28) |
| 15-24 y | 1.77*** (1.38-2.26) | 1.28*** (1.19-1.37) | 1.43*** (1.34-1.53) |
| **Education <sup>a</sup>** |  |  |  |
| No education | Ref. | Ref. | Ref. |
| Primary | 0.92 (0.76-1.11) | 0.76*** (0.72-0.80) | 0.83*** (0.78-0.88) |
| Secondary | 0.62*** (0.52-0.73) | 0.65*** (0.62-0.68) | 0.76*** (0.73-0.80) |
| Higher | 0.50*** (0.34-0.73) | 0.57*** (0.53-0.62) | 0.58*** (0.54-0.62) |
| **Mass media exposure <sup>b</sup>** |  |  |  |
| No | Ref. | Ref. | Ref. |
| Low | 1.07 (0.91-1.25) | 1.02 (0.97-1.07) | 0.96* (0.92-0.99) |
| High | 1.04 (0.86-1.25) | 0.83*** (0.78-0.88) | 0.90*** (0.84-0.95) |
| **Consumption of egg/fish/meat** |  |  |  |
| Never/occasionally | Ref. | Ref. | Ref. |
| At least once a week | 0.93 (0.82-1.06) | 0.77*** (0.74-0.80) | 0.91*** (0.88-0.94) |
| **Consumption of green leafy vegetables** |  |  |  |
| Never/occasionally | Ref. | Ref. | Ref. |
| Weekly | 1.18 (0.92-1.52) | 0.90*** (0.85-0.95) | 1.03 (0.97-1.09) |
| Daily | 1.15 (0.91-1.46) | 0.72*** (0.69-0.76) | 1.00 (0.95-1.06) |
| **ANC visits** |  |  |  |
| ≤ 4 ANC | Ref. | Ref. | Ref. |
| > 4 ANC | 1.17* (1.00-1.36) | 1.04 (1.00-1.08) | 1.16*** (1.12-1.20) |
| **Preceding birth interval** |  |  |  |
| ≤ 23 months | Ref. | Ref. | Ref. |
| > 23 months | 0.97 (0.76-1.23) | 1.01 (0.96-1.06) | 1.02 (0.96-1.08) |
| First birth | 1.11 (0.94-1.32) | 1.02 (0.96-1.09) | 1.06* (1.01-1.11) |
| **Postpartum haemorrhage** |  |  |  |
| No | Ref. | Ref. | Ref. |
| Yes | 1.06 (0.87-1.30) | 1.06* (1.01-1.10) | 1.09*** (1.05-1.13) |
| **100+ days IFA consumption** |  |  |  |
| No | Ref. | Ref. | Ref. |
| Yes | 1.25** (1.06-1.49) | 0.92*** (0.88-0.95) | 0.92*** (0.89-0.95) |
| **Received deworming** |  |  |  |
| No | Ref. | Ref. | Ref. |
| Yes | 1.05 (0.77-1.42) | 1.06** (1.01-1.11) | 1.08*** (1.04-1.12) |
| **Child's characteristics** |  |  |  |
| **Age** |  |  |  |
| 6-11 months | Ref. | Ref. | Ref. |
| 12-23 months | 1.07 (0.85-1.34) | 1.34*** (1.27-1.42) | 1.47*** (1.39-1.55) |
| 24-59 months | 0.80* (0.65-0.98) | 1.08** (1.02-1.14) | 1.19*** (1.13-1.25) |
| **Sex** |  |  |  |
| Male | Ref. | Ref. | Ref. |
| Female | 0.93 (0.82-1.06) | 0.99 (0.96-1.03) | 0.97* (0.94-0.99) |
| **Siblings under aged 5 years** |  |  |  |
| No | Ref. | Ref. | Ref. |
| Yes | 1.11 (0.93-1.33) | 1.18*** (1.13-1.24) | 1.16*** (1.11-1.21) |
|  | NFHS-3 (2006) | NFHS-4 (2016) | NFHS-5 (2021) |
| **Low birth weight/size** |  |  |  |
| No | Ref. | Ref. | Ref. |
| Yes | 1.16 (0.99-1.35) | 1.25*** (1.19-1.31) | 1.16*** (1.11-1.21) |
| **Received ICDS food supplementation** |  |  |  |
| No | Ref. | Ref. | Ref. |
| Yes | 0.80* (0.66-0.96) | 0.88*** (0.83-0.93) | 0.97 (0.91-1.04) |
| **Morbidity in last two weeks <sup>c</sup>** |  |  |  |
| No | Ref. | Ref. | Ref. |
| Yes | 1.03 (0.89-1.19) | 1.08*** (1.03-1.12) | 1.16*** (1.11-1.21) |
| **Household characteristics** |  |  |  |
| **Household size** |  |  |  |
| ≤5 members | Ref. | Ref. | Ref. |
| >5 members | 0.95 (0.83-1.08) | 1.11*** (1.07-1.15) | 1.00 (0.97-1.04) |
| **Place of residence** |  |  |  |
| Urban | Ref. | Ref. | Ref. |
| Rural | 0.78** (0.65-0.93) | 0.84*** (0.80-0.88) | 1.13*** (1.08-1.18) |
| **Caste** |  |  |  |
| Others | Ref. | Ref. | Ref. |
| Scheduled caste (SC)/Scheduled tribe (ST) | 0.97 (0.82-1.15) | 1.18*** (1.13-1.24) | 1.08*** (1.04-1.13) |
| Other backward classes (OBC) | 1.17 (0.98-1.39) | 1.12*** (1.07-1.18) | 0.87*** (0.84-0.91) |
| **Assets index** |  |  |  |
| Poor | Ref. | Ref. | Ref. |
| Middle | 0.74*** (0.63-0.87) | 0.96 (0.92-1.01) | 0.95* (0.91-0.99) |
| Rich | 0.65*** (0.51-0.83) | 1.03 (0.97-1.10) | 0.89*** (0.85-0.94) |
| **Piped drinking water <sup>d</sup>** |  |  |  |
| No | Ref. | Ref. | Ref. |
| Yes | 0.75*** (0.65-0.85) | 1.01 (0.97-1.05) | 0.98 (0.95-1.02) |
| **Toilet facility <sup>e</sup>** |  |  |  |
| Unimproved | Ref. | Ref. | Ref. |
| Improved | 1.01 (0.84-1.20) | 0.85*** (0.81-0.89) | 0.91*** (0.87-0.96) |
| **Community open defecation** | 2.96*** (2.35-3.72) | 1.76*** (1.65-1.88) | 1.61*** (1.49-1.74) |
| **Cooking fuel <sup>f</sup>** |  |  |  |
| Unclean | Ref. | Ref. | Ref. |
| Clean | 1.13 (0.92-1.40) | 1.05* (1.00-1.10) | 0.94*** (0.90-0.98) |

Level of significance \*\*\*p < 0.001, \*\*p < 0.01, \*p < 0.05;
<sup>a</sup>
Primary: schooling for 1-5 years, secondary: schooling for 6-12 years, higher: schooling for >12 years;
<sup>b</sup>
Mass media exposure was created by summing the frequency of exposure to the three media types: reading newspapers/magazines, listening to the radio, and watching TV and then categorizing them as 'no' for zero exposure, 'low' for exposure to one medium, and 'high' for exposure to more than one medium
<sup>c</sup>
Morbidity encompasses at least one of the three conditions: diarrhea, fever and acute respiratory infection (ARI)
<sup>d</sup>
Piped drinking water includes piped water, public taps/standpipes

### Projected reduction in anaemia among mother-child dyads under various scenarios of change in drivers

In a realistic scenario, i.e. if the trend from 2006 to 2021 continued at the same pace until 2030, the percentage of mothers with higher education would increase to 24% in 2030 (**Table 3, Panel A**). We estimate this increase in education would contribute to a 3.6% reduction in prevalence of anaemia among mother child dyads **(Table 3, Panel B)**. Similarly, an increase consistent with the prior trend in the percentage of households with improved toilet facilities (87% by 2030) would help eliminate 2.4% of anaemia burden. Between 2006 and 2021, the community open defecation dropped sharply, with an average annual rate of change (AARC) of −0.05. If this trend continues at the same pace until 2030, the prevalence of community open defecation is projected to drop to 11.4%, contributing to a 7.6% decrease in the prevalence of co-occurring anaemia among mother-child dyads. However, if the AARC improves to −0.08 (an optimistic scenario with a 3% increment in annual rate) from 2021 to 2030, the reduction in community open defecation prevalence would be even greater, leading to a further decline in anaemia prevalence. If the trends observed between 2006 and 2021 continue at a realistic rate, the combined effect of higher education, increased weekly consumption of eggs/fish/meat, daily consumption of green vegetables, 100+ days IFA consumption, ICDS food supplementation, improved toilet facilities, and reduced community open defecation predicts a 17.7% reduction in anaemia prevalence among mother-child dyads by 2030. Under more optimistic scenarios where improvements in risk factors accelerate, the predicted reduction in anaemia prevalence increases to 28.4%.

**Table 3:**
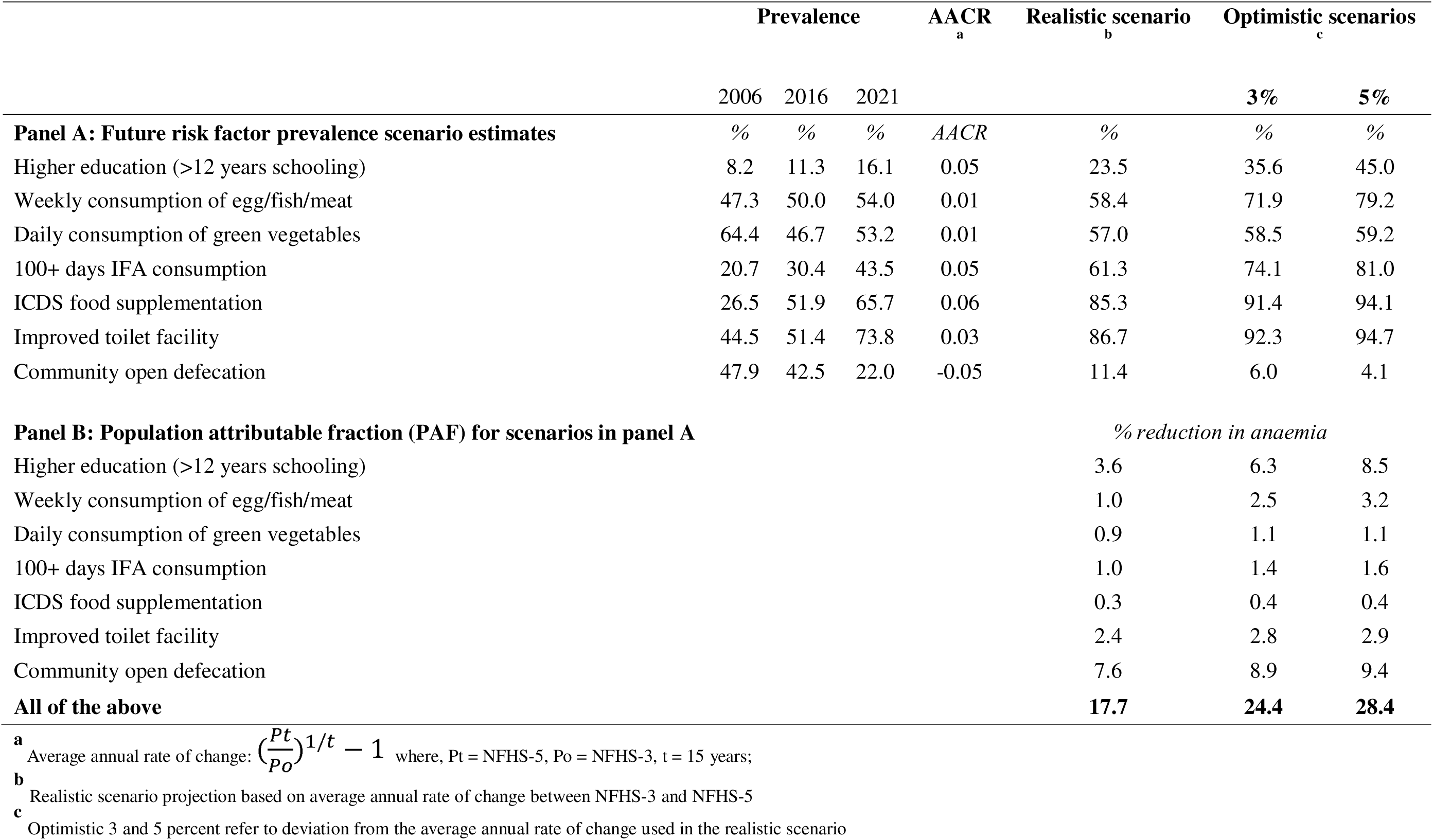
Projected reduction in anaemia among mother-child dyads (both anaemic) in India by 2030 under realistic and optimistic scenarios.

## Discussion

We studied anaemia in over 400,000 mother-child dyads from 2006 to 2021 in India. In 2021, at least one member of the dyad was anaemic in 78% of dyads, and both members were anaemic in 37% of dyads. Overall, the prevalence of anaemia among mother-child dyads has not changed substantially over time, though trends vary by state or district. We further classified dyads by anaemia severity and found that moderate/severe cases (category 4) increased from 2016 to 2021 whereas mild cases (category 2) decreased during this period. Maternal education, animal source food consumption, and healthcare utilization increased over time and were associated with lower odds of anaemia in these dyads while community-level open defecation was associated with increased odds. Our forward-looking analysis suggests that accelerating improvements in maternal education, weekly consumption of egg/fish/meat, and sanitation facilities, and reducing community open defecation could contribute to an 18% to 28% reduction in anaemia prevalence among mother-child dyads in India by 2030, suggesting that factors other than those measured in NFHS should be addressed for total elimination of this persistent public health issue.

In accordance with Biradar et al. (2023), we found a high prevalence of anaemia among mother-child dyads in India. Moreover, our study found that 12.6% of mother-child dyads were affected by severe/moderate anaemia. This high prevalence underscores a significant public health concern. The potential reasons for this include a shared unhealthy home environment, poor dietary intake, and limited access to healthcare (WHO, 2017). We also found variation in anaemia prevalence among mother-child pairs by state or district, with some states/districts showing improvements and others worsening. Explaining this variation was not an objective of the current analysis, but drivers of subnational variation need to be better understood to tailor anaemia reduction strategies to specific contexts. Such an effort is likely highly complex given the broad set of determinants of anaemia across individual, household, community, social, and political levels.

Our findings on which factors were associated with anaemia among mother-child dyads concur with previous research on factors associated with anaemia in individuals. Low maternal education has been associated with anaemia in women (Wiafe et al., 2023; Sharma et al., 2024) and children (Gebreegziabher & Sidibe, 2023; Onyeneho et al., 2019; Choi et al., 2011). Education may provide mothers with the knowledge, resources, and empowerment to prevent and manage anaemia effectively. We also found an association between media exposure and anaemia, which is consistent with previous studies (Armah-Ansah, 2023; Mbule et al., 2013) and suggests connectedness to sources of information, potentially on healthy diets or health services for example, may be beneficial. The association we found between animal source food consumption and anaemia has also been documented (Saaka & Rauf, 2015; Grover et al., 2020). These foods are good sources of heme iron (Moustarah & Daley, 2022), which is absorbed better than non-heme iron found in plant sources.

Several factors related to the health system were associated with anaemia in mother-child dyads. Postpartum haemorrhage can deplete the mother’s iron stores, limiting haemoglobin production (Carroli et al., 2008). In some cases, postpartum follow-up and nutritional support may be inadequate, leading to prolonged anaemia in mothers, which may then affect the child’s health. This finding underscores the importance of institutional deliveries, the quality of care during delivery, and complication management. We also found a significant association between IFA consumption and anaemia among mother-child dyads, in line with studies by Sartika et al. (2024) and Palika et al. (2022). IFA supplements help replenish the body’s iron stores, which are crucial for haemoglobin production (WHO, 2017; Abbaspour et al., 2014). Infants born to mothers with adequate iron levels are more likely to have sufficient iron stores at birth, reducing the risk of anaemia in early infancy (Shukla et al., 2019).

We found higher odds of anaemia among dyads where children had siblings under five years of age. In households with multiple young children, resources must be spread out and this has consequences for outcomes such as dietary intake and receipt of health services. The utilization of ICDS, which provides essential nutrients such as iron, folic acid, and vitamins through fortified take-home rations and hot-cooked meals, has been linked to reduced odds of anaemia among children in India (Hirve et al., 2013). Community-based delivery of the ICDS transfers through Anganwadi centers ensures consistent access to these supplements.

Consistent with the finding of Biradar (2023) and Khan et al. (2020), we found that the coexistence of anaemia among mother-child dyads was lower in wealthy households, which may reflect better access to nutritious foods, superior healthcare services, awareness about health and nutrition, and a more hygienic residential environment. Women and children in poor households often lack access to preventive and therapeutic supplements, such as vitamins and iron tablets (WHO, 2017). Enhancing living conditions and sanitation may lower the risk of infections that can lead to anaemia. Community open defecation can contaminate the environment, elevating the risk of faecal-oral transmission of diseases caused by intestinal helminths, protozoans, bacteria, and viruses (Hutton & Chase, 2017). These organisms can cause intestinal bleeding and nutrient malabsorption, which deplete iron levels in the body (WHO, 2023). Additionally, the increased exposure to pathogens can result in chronic infections that further exacerbate iron deficiency and anaemia. Improved sanitation practices, including eliminating open defecation, are crucial in reducing these health risks and preventing anaemia.

The projection and PAF estimates highlight the importance of a holistic approach in reducing anaemia among mother-child dyads in India. In the last fifteen years, there have been substantial improvements in women’s higher education, IFA consumption, ICDS food supplementation, and sanitation. Future education and community sanitation improvements showed the most potential for reducing anaemia, as evidenced by their high PAF estimates. Improving diets – consumption of animal source foods and green leafy vegetables – can also contribute. The cumulative PAF suggests that, if the seven studied risk factors continue to improve at the same rate as they have since 2006, the co-occurrence of anaemia among mother-child dyads would reduce to around 19% by 2030, from a baseline of 37% in 2021. In more optimistic scenarios, where improvements in these risk factors accelerate, anaemia prevalence could potentially fall even further, highlighting the significant impact of ambitious, well-coordinated public health interventions. Thus, enhancing maternal education, dietary diversity, iron supplementation and sanitation remain pivotal in combating anaemia, necessitating sustained and integrated public health efforts.

Our findings particularly PAF should be interpreted with caution since the use of cross-sectional data in the analysis prevents establishing causal inferences. Any projections based on repeated cross-sectional data will inherently involve some degree of uncertainty. Further, the analysis was confined to factors available in the NFHS, leaving out potentially important factors such as genetic haemoglobin disorders, deficiencies of vitamin A, vitamin B12, infectious diseases, malaria, and HIV/AIDS. This introduces the possibility of unrestricted confounding factors not accounted for in the model. Finally, the use of the capillary blood samples instead of venous blood may result in inaccurate estimation of anaemia (Hackl et al., 2024). NFHS and other DHS surveys use capillary blood due to its operational feasibility, affordability, and user-friendliness.

## Conclusion

We provide new evidence on the co-occurrence of anaemia among mother-child dyads at national and subnational levels, taking a forward-looking approach to understand opportunities to address this persistent public health problem. Our findings highlight an increase in co-occurrence of anaemia in these dyads from 2016 to 2021. To effectively reduce anaemia prevalence by 2030, a goal being actively pursued through the nationwide Anemia Mukt Bharat (Anemia Free India) program under the Ministry of Health and Family Welfare, our study findings support targeted interventions to increase maternal education, improve sanitation facilities, reduce open defecation, and promote consumption of nutritious foods. Sustained and integrated public health efforts focusing on these areas are recommended to combat anaemia effectively and ensure better health for mothers and children across India.

## Author contributions

SP conceptualized the manuscript, was involved in the statistical analysis, and reviewed the manuscript. TM performed the statistical analysis, prepared the tables and figures, wrote the initial draft, and revised the manuscript. MG contributed to data visualization and reviewed the results for accuracy. SC led the statistical analysis and reviewed the manuscript. PHN, SKS, RA, and LKD provided critical reviews and contributed to writing the manuscript. SS reviewed and edited the manuscript and provided intellectual insights. All authors read and approved the final submitted version.

## Data availability

The data that support the findings of this study are available online on the website of the Demographic Health Survey (DHS): https://dhsprogram.com/Countries/Country-Main.cfm?ctry_id=57&c=India&Country=India&cn=&r=4.

## Ethics statement

All documents and data sources used and analysed in this study are already in the public domain and freely available. Sources of the original documents and data have been acknowledged and cited.

