## Supplementary material for "Anaemia among mother-child dyads in India: trends, drivers, and future projections": Table S

**Table S1: Trends in anaemia among mother-child dyads (both anaemic) by states/UTs, 2006, 2016, 2021**

| **State/UT** | **Both anaemic** | | |
| --- | --- | --- | --- |
|  | **2006 (%)** | **2016 (%)** | **2021 (%)** |
| **North** | **32.8** | **34.4** | **37.5** |
| Chandigarh | - | 57.1 | 28.8 |
| Delhi | 16.9 | 30.5 | 32.1 |
| Haryana | 41.9 | 47.5 | 41.2 |
| Himachal Pradesh | 24.3 | 29.7 | 28.3 |
| Jammu Kashmir | 31.9 | 28.3 | 45.9 |
| Punjab | 28.5 | 31.6 | 39.5 |
| Rajasthan | 36.9 | 32.4 | 38.4 |
| Uttarakhand | 31.3 | 30.3 | 23.3 |
| **Central** | **36.8** | **35.5** | **34.7** |
| Chhattisgarh | 38.1 | 23.4 | 40.1 |
| Madhya Pradesh | 42.6 | 38.3 | 38.5 |
| Uttar Pradesh | 33.8 | 36.1 | 32.8 |
| **East** | **42.2** | **36.8** | **43.7** |
| Bihar | 51.5 | 39.0 | 42.7 |
| Jharkhand | 45.2 | 48.3 | 44.6 |
| Odisha | 34.9 | 25.9 | 40.5 |
| West Bengal | 38.1 | 33.6 | 46.5 |
| **Northeast** | **28.9** | **17.3** | **37.4** |
| Arunachal Pradesh | 33.9 | 22.8 | 24.5 |
| Assam | 44.6 | 16.6 | 42.6 |
| Manipur | 16.6 | 8.3 | 15.7 |
| Meghalaya | 26.6 | 26.9 | 23.8 |
| Mizoram | 20.0 | 7.1 | 17.7 |
| Nagaland | 0.0 | 9.7 | 13.7 |
| Sikkim | 29.1 | 21.0 | 16.4 |
| Tripura | 41.4 | 25.4 | 40.3 |
| **West** | **39.0** | **29.5** | **40.1** |
| DNH and DD | - | 58.2 | 45.2 |
| Goa | 12.9 | 19.1 | 19.9 |
| Gujarat | 39.0 | 35.2 | 50.5 |
| Maharashtra | 26.4 | 26.7 | 35.1 |
| **South** | **31.2** | **27.1** | **28.5** |
| Andaman and Nicobar Islands | - | 30.6 | 22.7 |
| Andhra Pradesh | 41.4 | 29.8 | 34.3 |
| Karnataka | 24.5 | 27.0 | 29.5 |
| Kerala | 14.0 | 13.7 | 13.5 |
| Lakshadweep | - | 25.0 | 11.3 |
| Puducherry | - | 25.8 | 30.8 |
| Tamil Nadu | 33.5 | 27.7 | 28.1 |
| Telangana | - | 31.4 | 33.0 |
| **India** | 35.0 | **32.6** | **37.1** |

**Table S2: Trend in anaemia among different categories of severity in mother-child pair by states/UTs, 2006, 2016, 2021**

| **State/UT** | **Category 2** | | | **Category 3** | | | **Category 4** | | |
| --- | --- | --- | --- | --- | --- | --- | --- | --- | --- |
|  | **2006 (%)** | **2016 (%)** | **2021 (%)** | **2006 (%)** | **2016 (%)** | **2021 (%)** | **2006 (%)** | **2016 (%)** | **2021 (%)** |
| **North** | **23.1** | **28.2** | **26.7** | **22.9** | **20.8** | **21.5** | **27.9** | **27.8** | **30.5** |
| Chandigarh | - | 20.1 | 30.2 | - | 16.8 | 19.5 | - | 51.1 | 16.5 |
| Delhi | 19.8 | 27.6 | 25.6 | 16.6 | 21.9 | 21.9 | 13.6 | 26.2 | 26.5 |
| Haryana | 22.9 | 25.2 | 24.6 | 22.8 | 23.4 | 21.4 | 35.8 | 39.7 | 34.8 |
| Himachal Pradesh | 25.9 | 28.1 | 31.8 | 19.0 | 19.4 | 18.9 | 18.9 | 22.8 | 22.1 |
| Jammu Kashmir | 26.5 | 25.2 | 21.8 | 20.1 | 23.0 | 24.0 | 25.3 | 23.4 | 40.2 |
| Punjab | 23.9 | 31.0 | 23.4 | 24.4 | 21.5 | 21.2 | 24.3 | 24.8 | 32.9 |
| Rajasthan | 21.2 | 29.1 | 28.8 | 25.8 | 19.2 | 21.5 | 32.4 | 25.7 | 30.5 |
| Uttarakhand | 29.2 | 28.7 | 24.3 | 19.6 | 21.8 | 21.0 | 24.7 | 23.8 | 18.0 |
| **Central** | 24.2 | **28.6** | **27.0** | 22.6 | **20.9** | **21.4** | 30.8 | **28.3** | **28.1** |
| Chhattisgarh | 26.9 | 31.2 | 28.2 | 23.1 | 16.8 | 20.9 | 31.1 | 16.7 | 32.5 |
| Madhya Pradesh | 29.7 | 28.9 | 25.7 | 19.2 | 21.7 | 21.4 | 34.5 | 30.6 | 31.5 |
| Uttar Pradesh | 21.1 | 28.1 | 27.2 | 24.1 | 21.2 | 21.4 | 28.9 | 29.1 | 26.5 |
| **East** | 30.5 | **31.9** | **29.9** | 17.9 | **19.3** | **19.2** | 32.7 | **28.1** | **34.3** |
| Bihar | 30.2 | 31.1 | 29.3 | 16.3 | 20.5 | 19.6 | 40.0 | 30.1 | 34.0 |
| Jharkhand | 26.7 | 29.8 | 28.1 | 18.4 | 17.1 | 18.9 | 34.9 | 37.9 | 35.4 |
| Odisha | 32.0 | 31.4 | 30.8 | 18.4 | 15.4 | 19.6 | 26.6 | 19.1 | 31.0 |
| West Bengal | 30.9 | 34.4 | 31.2 | 18.6 | 19.9 | 18.4 | 29.6 | 24.8 | 35.8 |
| **Northeast** | 22.0 | **31.5** | **31.8** | 22.0 | **17.5** | **18.9** | 24.6 | **11.8** | **28.6** |
| Arunachal Pradesh | 25.2 | 31.2 | 31.0 | 19.4 | 18.6 | 18.4 | 27.6 | 16.7 | 17.9 |
| Assam | 20.9 | 32.6 | 32.7 | 22.9 | 17.5 | 18.9 | 38.8 | 11.1 | 32.9 |
| Manipur | 34.2 | 26.1 | 31.7 | 14.8 | 13.1 | 13.7 | 10.2 | 4.8 | 9.9 |
| Meghalaya | 25.7 | 30.1 | 26.4 | 22.0 | 22.8 | 22.3 | 21.3 | 20.1 | 18.5 |
| Mizoram | 27.8 | 20.4 | 30.8 | 18.7 | 11.7 | 17.1 | 16.1 | 4.7 | 12.0 |
| Nagaland | 0.0 | 22.8 | 29.7 | 0.0 | 15.1 | 15.0 | 0.0 | 7.1 | 8.5 |
| Sikkim | 28.9 | 35.7 | 27.6 | 23.9 | 15.7 | 19.0 | 22.4 | 13.8 | 11.8 |
| Tripura | 30.9 | 34.0 | 32.4 | 18.6 | 18.2 | 19.8 | 32.5 | 17.6 | 30.8 |
| **West** | 24.0 | **31.9** | **25.9** | 24.3 | **19.3** | **20.9** | 32.5 | **22.6** | **32.8** |
| DNH and DD | - | 25.3 | 30.9 | - | 17.8 | 15.5 | - | 46.5 | 37.0 |
| Goa | 23.1 | 33.5 | 31.2 | 16.1 | 20.1 | 20.8 | 9.7 | 11.1 | 14.4 |
| Gujarat | 24.0 | 31.4 | 23.4 | 24.3 | 20.2 | 20.8 | 32.5 | 26.9 | 43.0 |
| Maharashtra | 21.7 | 32.1 | 27.0 | 22.8 | 18.8 | 21.0 | 22.6 | 20.5 | 27.8 |
| **South** | 27.1 | **29.8** | **27.5** | 21.7 | **21.2** | **20.3** | 25.3 | **21.1** | **22.6** |
| Andaman and Nicobar Islands | - | 27.0 | 36.4 | - | 23.4 | 16.2 | - | 26.0 | 16.8 |
| Andhra Pradesh | 26.1 | 27.1 | 27.0 | 22.0 | 23.2 | 21.4 | 35.0 | 24.8 | 27.2 |
| Karnataka | 24.7 | 31.0 | 27.3 | 24.0 | 21.6 | 21.1 | 19.8 | 20.7 | 23.6 |
| Kerala | 28.3 | 31.3 | 30.3 | 17.6 | 13.6 | 12.6 | 10.2 | 8.2 | 9.0 |
| Lakshadweep | - | 34.9 | 34.2 | - | 17.6 | 16.5 | - | 16.3 | 3.7 |
| Puducherry | - | 35.9 | 28.8 | - | 18.7 | 24.7 | - | 14.6 | 23.8 |
| Tamil Nadu | 30.4 | 31.4 | 29.0 | 21.3 | 22.2 | 20.9 | 25.8 | 20.7 | 21.8 |
| Telangana | - | 26.9 | 22.3 | - | 21.1 | 22.7 | - | 26.8 | 28.5 |
| **India** | **25.8** | **30.2** | **27.8** | **21.4** | **20.2** | **20.5** | **28.6** | **25.5** | **29.7** |

**_Note:_** _Category 2:_ _mild and non-anaemic dyad; Category 3: severe/moderate and non-anaemic; Category 4: mild and moderate/severely anaemic dyad_

**Figure S1: Prevalence of anaemia among mother-child pair by districts, 2016, 2021**

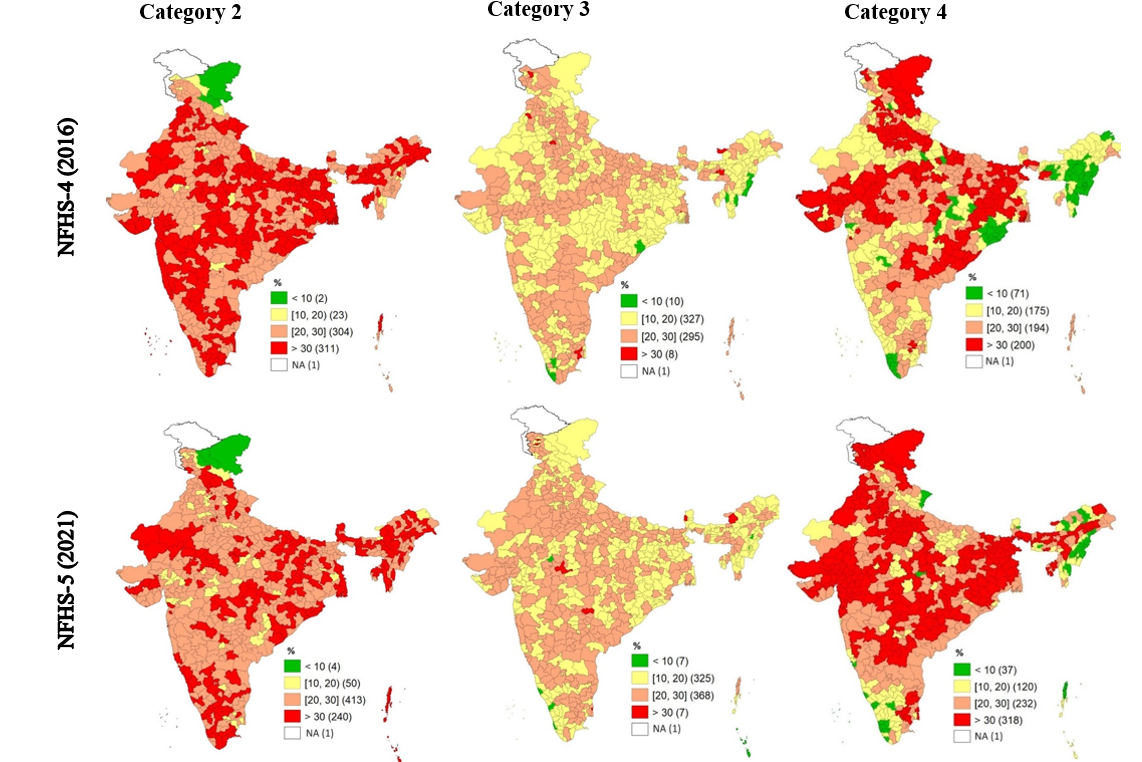

**_Note:_** _NFHS-3 does not provide district level data;_

_Category 2:_ _mild and non-anaemic dyad; Category 3: severe/moderate and non-anaemic; Category 4: mild and moderate/severely anaemic dyad_

**Table S3: Absolute and relative changes in the background characteristics, 2006, 2016, 2021**

| **Background characteristics** | **Absolute Change (pp) (NFHS 3 & NFHS 4) ^a^** | **Absolute Change (pp) (NFHS 4 & NFHS 5)** | **Relative change (%) (NFHS 3 & 4) ^b^** | **Relative change (%) (NFHS 4 & 5)** |
| --- | --- | --- | --- | --- |
| **Mother’s characteristics** | | | | |
| Age (15-24 years) | -8.7*** | -2.3*** | -21.2 | -7.3 |
| Age (25-34 years) | 6.8*** | 2.3*** | 13.2 | 4.0 |
| Age (35-49 years) | 1.9*** | 0.0 | 26.6 | 0.1 |
| Higher education (>12 years schooling) | 2.8*** | 5.1*** | 34.1 | 46.4 |
| Low media exposure **^c^** | 9.1*** | -4.3*** | 25.5 | -9.7 |
| High media exposure | -7.9*** | -11.8** | -26.2 | -53.6 |
| At least weekly consumption of egg/fish/meat | 2.7*** | 4.0*** | 5.7 | 8.0 |
| Weekly consumption of green leafy vegetables | 9.8*** | -0.6 | 34.0 | -1.6 |
| Daily consumption of green leafy vegetables | -17.7*** | 6.5*** | -27.5 | 13.9 |
| 4+ ANC visits | 0.9 | 1.9*** | 2.3 | 4.7 |
| > 23 months preceding birth interval | -1.6*** | -0.4 | -3.3 | -1.1 |
| Postpartum haemorrhage | 6.8*** | 3.5** | 53.5 | 18.5 |
| 100+ days IFA consumption | 9.8*** | 12.9*** | 46.9 | 43.1 |
| Received Deworming | 13.8*** | 12.9*** | 318.2 | 71.2 |
| **Child's characteristics** | | | | |
| Age (6-11 months) | -1.1*** | -0.6*** | -8.1 | -5.6 |
| Age (12-23 months) | -0.7 | -0.4* | -3.1 | -1.4 |
| Age (24-59 months) | 1.8*** | 1.0*** | 2.8 | 1.5 |
| Male | -0.9* | -0.3 | -1.7 | -0.8 |
| Female | 0.9* | 0.3 | 2.0 | 0.9 |
| Siblings <5 years | -2.3*** | -1.8*** | -5.4 | -4.5 |
| Low birth weight/size | -3.8*** | 0.2 | -18.8 | 1.2 |
| ICDS food supplementation | 25.2*** | 14.0*** | 95.1 | 27.1 |
| Morbidity in last 2 weeks **^d^** | -2.8*** | -0.7** | -13.0 | -3.7 |
| **Household characteristics** | | | | |
| >5 household size | -0.3 | -1.6*** | -0.5 | -3.0 |
| Rural | 4.1** | 1.7*** | 6.2 | 2.6 |
| Scheduled caste (SC)/Scheduled tribe (ST) | 3.6*** | 1.2** | 9.3 | 3.8 |
| Other backward classes (OBC) | 7.0*** | -0.6 | 15.9 | -1.4 |
| Asset index (poor) | -13.5*** | -1.1** | -27.4 | -3.0 |
| Asset index (middle) | 0.9 | 5.0*** | 3.3 | 18.0 |
| Asset index (rich) | 12.6*** | -3.8*** | 53.6 | -10.8 |
| Pipped drinking water **^e^** | -2.7** | 2.5*** | -8.5 | 5.8 |
| Improved toilet facility **^f^** | 7.4*** | 21.7*** | 15.5 | 43.6 |
| Clean cooking fuel **^h^** | 9.6** | 14.7*** | 36.5 | 41.6 |

**^a^** _Absolute change = NFHS4-NFHS3_

**^b^** _Relative change_ _= ((NFHS4-NFHS3)/NFHS3)*100_

**^c^** _Mass media exposure was created by summing the frequency of exposure to the three media types: reading newspapers/magazines, listening to the radio, and watching TV and then categorizing them as 'no' for zero exposure, 'low' for exposure to one medium, and 'high' for exposure to more than one medium_

**^d^** _Morbidity encompasses at least one of the three conditions: diarrhea, fever and acute respiratory infection (ARI)_

**^e^** _Piped drinking water includes piped water, public taps/standpipes_

**^f^** _Improved toilet facilities include any non-shared toilet of the following types: flush/pour flush toilets to piped sewer systems, septic tanks, pit latrines, or an unknown destination; ventilated improved pit (VIP)/biogas latrines; pit latrines with slabs; and twin pit/composting toilets_

**^g^** _Community open defecation was measured as the average proportion of households that reported practising open defecation in a primary sampling unit_

**^h^** _Clean cooking fuel includes electricity, LPG/natural gas, biogas_

_Indicators such as ANC visits, preceding birth interval, 100+ days IFA consumption, received deworming are for the last birth_

**Table S4: Association between selected factors and anaemia among mother-child dyads: Results from regression analysis**

| **Background characteristics** | **AOR (95% CI)** | | | | | | | | |
| --- | --- | --- | --- | --- | --- | --- | --- | --- | --- |
|  | **Category 2** | | | **Category 3** | | | **Category 4** | | |
|  | **2006** | **2016** | **2021** | **2006** | **2016** | **2021** | **2006** | **2016** | **2021** |
| **Mother's characteristics** | | | | | | | | | |
| **Age (in yr.)** |  |  |  |  |  |  |  |  |  |
| 35 & above | Ref. | Ref. | Ref. | Ref. | Ref. | Ref. | Ref. | Ref. | Ref. |
| 25-34 | 1.38**(1.10-1.73) | 1.08*(1.02-1.15) | 1.14***(1.08-1.21) | 1.34*(1.05-1.72) | 1.02 (0.96-1.09) | 1.11***(1.04-1.18) | 1.36**(1.08-1.73) | 1.08*(1.01-1.15) | 1.20***(1.13-1.27) |
| 15-24 | 1.79***(1.39-2.32) | 1.13***(1.06-1.21) | 1.25***(1.17-1.34) | 1.51**(1.15-2.00) | 1.07 (0.99-1.15) | 1.23***(1.13-1.32) | 1.71***(1.31-2.22) | 1.26***(1.17-1.36) | 1.43***(1.34-1.53) |
| **Education ^a^** |  |  |  |  |  |  |  |  |  |
| No education | Ref. | Ref. | Ref. | Ref. | Ref. | Ref. | Ref. | Ref. | Ref. |
| Primary | 0.86 (0.70-1.05) | 0.89***(0.84-0.94) | 0.91**(0.86-0.97) | 0.85 (0.68-1.05) | 0.88***(0.83-0.94) | 0.84***(0.79-0.90) | 0.97 (0.79-1.18) | 0.75***(0.71-0.80) | 0.82***(0.77-0.87) |
| Secondary | 0.71***(0.59-0.85) | 0.81***(0.77-0.85) | 0.88***(0.84-0.93) | 0.63***(0.52-0.76) | 0.75***(0.71-0.79) | 0.79***(0.75-0.83) | 0.62***(0.52-0.74) | 0.63***(0.60-0.66) | 0.75***(0.71-0.79) |
| Higher | 0.62**(0.43-0.89) | 0.81***(0.75-0.87) | 0.79***(0.73-0.84) | 0.55**(0.36-0.83) | 0.70***(0.64-0.76) | 0.65***(0.61-0.70) | 0.50***(0.33-0.75) | 0.55***(0.51-0.60) | 0.55***(0.52-0.59) |
| **Mass media exposure ^b^** |  |  |  |  |  |  |  |  |  |
| No | Ref. | Ref. | Ref. | Ref. | Ref. | Ref. | Ref. | Ref. | Ref. |
| Low | 0.97 (0.82-1.15) | 0.99 (0.95-1.04) | 0.96*(0.92-0.99) | 1.15 (0.96-1.39) | 1.02 (0.97-1.07) | 0.98 (0.94-1.02) | 1.07 (0.91-1.27) | 1.04 (1.00-1.10) | 0.96 (0.92-1.00) |
| High | 1.19 (0.98-1.45) | 0.88***(0.82-0.93) | 0.90**(0.85-0.96) | 1.07 (0.86-1.33) | 0.90**(0.84-0.96) | 0.91**(0.85-0.97) | 1.04 (0.85-1.27) | 0.85***(0.80-0.91) | 0.92*(0.86-0.98) |
| **Consumption of animal food** |  |  |  |  |  |  |  |  |  |
| Never/occasionally | Ref. | Ref. | Ref. | Ref. | Ref. | Ref. | Ref. | Ref. | Ref. |
| At least once a week | 1.039 (0.91-1.19) | 0.92***(0.88-0.95) | 1.00 (0.94-1.07) | 0.87 (0.75-1.01) | 0.85***(0.82-0.88) | 0.91***(0.87-0.94) | 0.91 (0.79-1.05) | 0.75***(0.72-0.78) | 0.89***(0.86-0.92) |
| **Consumption of green leafy vegetables** |  |  |  |  |  |  |  |  |  |
| Never/occasionally | Ref. | Ref. | Ref. | Ref. | Ref. | Ref. | Ref. | Ref. | Ref. |
| Weekly | 0.95 (0.74-1.23) | 0.97 (0.92-1.03) | 1.00 (0.94-1.07) | 1.28 (0.95-1.73) | 0.93*(0.88-0.99) | 1.01 (0.94-1.08) | 1.19 (0.92-1.55) | 0.87***(0.82-0.92) | 1.01 (0.95-1.07) |
| Daily | 1.04 (0.82-1.33) | 0.88***(0.83-0.93) | 1.04 (0.98-1.10) | 1.42* (1.07-1.88) | 0.83***(0.78-0.88) | 1.01 (0.95-1.08) | 1.14 (0.89-1.47) | 0.69***(0.65-0.73) | 0.97 (0.91-1.03) |
| **ANC visits** |  |  |  |  |  |  |  |  |  |
| ≤ 4 ANC | Ref. | Ref. | Ref. | Ref. | Ref. | Ref. | Ref. | Ref. | Ref. |
| > 4 ANC | 1.29**(1.11-1.51) | 1.02 (0.99-1.06) | 1.07***(1.03-1.11) | 1.08 (0.91-1.29) | 1.07**(1.03-1.12) | 1.09***(1.05-1.13) | 1.13 (0.96-1.33) | 1.04 (1.00-1.08) | 1.17***(1.13-1.22) |
| **Preceding birth interval** |  |  |  |  |  |  |  |  |  |
| ≤ 23 months | Ref. | Ref. | Ref. | Ref. | Ref. | Ref. | Ref. | Ref. | Ref. |
| > 23 months | 0.963 (0.75-1.24) | 1.04 (0.97-1.12) | 1.06 (0.99-1.13) | 0.88 (0.67-1.15) | 1.00 (0.92-1.07) | 1.02 (0.97-1.08) | 0.99 (0.76-1.28) | 1.0 (0.9-1.08) | 1.01 (0.94-1.07) |
| First birth | 1.29**(1.07-1.55) | 1.04 (0.98-1.09) | 1.09***(1.04-1.15) | 1.08 (0.89-1.32) | 0.99 (0.93-1.05) | 1.05 (0.98-1.13) | 1.08 (0.90-1.30) | 0.98 (0.93-1.04) | 1.06*(1.01-1.11) |
| **Postpartum haemorrhage** |  |  |  |  |  |  |  |  |  |
| No | Ref. | Ref. | Ref. | Ref. | Ref. | Ref. | Ref. | Ref. | Ref. |
| Yes | 1.08 (0.88-1.32) | 1.03 (0.98-1.08) | 1.02 (0.98-1.06) | 1.29*(1.04-1.60) | 1.05* (1.00-1.10) | 1.06**(1.02-1.11) | 1.06 (0.86-1.31) | 1.08**(1.03-1.13) | 1.09***(1.05-1.14) |
| **100+ days iron tablet consumption** |  |  |  |  |  |  |  |  |  |
| No | Ref. | Ref. | Ref. | Ref. | Ref. | Ref. | Ref. | Ref. | Ref. |
| Yes | 1.22* (1.02-1.45) | 0.97 (0.94-1.01) | 0.96*(0.92-0.99) | 1.3** (1.07-1.58) | 0.92***(0.88-0.96) | 0.94***(0.90-0.97) | 1.22*(1.02-1.47) | 0.91***(0.87-0.95) | 0.90***(0.87-0.94) |
| **Received deworming** |  |  |  |  |  |  |  |  |  |
| No | Ref. | Ref. | Ref. | Ref. | Ref. | Ref. | Ref. | Ref. | Ref. |
| Yes | 0.95 (0.69-1.30) | 1.14***(1.09-1.19) | 1.07***(1.04-1.11) | 1.19 (0.85-1.65) | 1.06**(1.02-1.12) | 1.07***(1.03-1.11) | 0.99 (0.72-1.37) | 1.04 (0.99-1.09) | 1.08***(1.04-1.12) |
| **Child's characteristics** | | | | | | | | | |
| **Age** |  |  |  |  |  |  |  |  |  |
| 6-11 months | Ref. | Ref. | Ref. | Ref. | Ref. | Ref. | Ref. | Ref. | Ref. |
| 12-23 months | 0.89 (0.70-1.13) | 1.09**(1.03-1.16) | 1.20***(1.14-1.28) | 0.95 (0.73-1.24) | 1.06 (1.00-1.14) | 1.21***(1.14-1.29) | 1.15 (0.90-1.46) | 1.40***(1.32-1.49) | 1.48***(1.40-1.57) |
| 24-59 months | 0.83 (0.68-1.03) | 0.98 (0.93-1.03) | 1.07**(1.02-1.14) | 0.81 (0.64-1.02) | 0.92**(0.87-0.98) | 0.99 (0.94-1.05) | 0.81 (0.66-1.01) | 1.09**(1.03-1.16) | 1.18***(1.12-1.24) |
| **Sex** |  |  |  |  |  |  |  |  |  |
| Male | Ref. | Ref. | Ref. | Ref. | Ref. | Ref. | Ref. | Ref. | Ref. |
| Female | 1.00 (0.88-1.14) | 1.00 (0.97-1.04) | 0.98 (0.94-1.01) | 0.92 (0.79-1.06) | 0.98 (0.94-1.02) | 0.98 (0.94-1.01) | 0.94 (0.82-1.07) | 0.98 (0.94-1.01) | 0.96**(0.93-0.99) |
| **Siblings under aged 5** |  |  |  |  |  |  |  |  |  |
| No | Ref. | Ref. | Ref. | Ref. | Ref. | Ref. | Ref. | Ref. | Ref. |
| Yes | 1.05 (0.88-1.26) | 1.07**(1.02-1.13) | 1.07**(1.02-1.12) | 0.89 (0.73-1.08) | 1.12***(1.06-1.18) | 1.18***(1.13-1.25) | 1.11 (0.92-1.34) | 1.21***(1.15-1.28) | 1.18***(1.13-1.24) |
| **Low birth weight/size** |  |  |  |  |  |  |  |  |  |
| No | Ref. | Ref. | Ref. | Ref. | Ref. | Ref. | Ref. | Ref. | Ref. |
| Yes | 0.98 (0.83-1.15) | 1.08**(1.03-1.13) | 1.03 (0.98-1.08) | 1.09 (0.91-1.30) | 1.2***(1.13-1.26) | 1.13***(1.08-1.19) | 1.21*(1.03-1.42) | 1.29***(1.23-1.35) | 1.20***(1.15-1.26) |
| **ICDS food supplementation** |  |  |  |  |  |  |  |  |  |
| No | Ref. | Ref. | Ref. | Ref. | Ref. | Ref. | Ref. | Ref. | Ref. |
| Yes | 0.74**(0.61-0.90) | 0.91**(0.86-0.97) | 0.99 (0.93-1.07) | 0.74**(0.60-0.91) | 0.93 (0.87-1.00) | 0.99 (0.92-1.07) | 0.80*(0.65-0.97) | 0.87***(0.81-0.92) | 0.96 (0.89-1.03) |
| **Morbidity in last two weeks ^c^** |  |  |  |  |  |  |  |  |  |
| No | Ref. | Ref. | Ref. | Ref. | Ref. | Ref. | Ref. | Ref. | Ref. |
| Yes | 1.01 (0.87-1.18) | 1.00 (0.96-1.05) | 1.09***(1.04-1.14) | 1.02 (0.87-1.21) | 1.04 (0.99-1.09) | 1.12***(1.07-1.17) | 1.05 (0.90-1.23) | 1.09***(1.04-1.14) | 1.18***(1.13-1.23) |
| **Household characteristics** | | | | | | | | | |
| **Household size** |  |  |  |  |  |  |  |  |  |
| ≤5 members | Ref. | Ref. | Ref. | Ref. | Ref. | Ref. | Ref. | Ref. | Ref. |
| >5 members | 0.99 (0.87-1.14) | 1.05*(1.01-1.08) | 1.01 (0.97-1.04) | 0.99 (0.86-1.16) | 1.07***(1.03-1.11) | 1.00 (0.96-1.04) | 0.93 (0.81-1.07) | 1.12***(1.08-1.16) | 1.01 (0.98-1.05) |
| **Place of residence** |  |  |  |  |  |  |  |  |  |
| Urban | Ref. | Ref. | Ref. | Ref. | Ref. | Ref. | Ref. | Ref. | Ref. |
| Rural | 0.92 (0.77-1.11) | 0.90***(0.86-0.95) | 1.08***(1.04-1.14) | 0.97 (0.80-1.19) | 0.88***(0.83-0.93) | 1.08**(1.03-1.14) | 0.80*(0.66-0.96) | 0.84***(0.79-0.88) | 1.14***(1.09-1.19) |
| **Caste** |  |  |  |  |  |  |  |  |  |
| Others | Ref. | Ref. | Ref. | Ref. | Ref. | Ref. | Ref. | Ref. | Ref. |
| SC/ST | 0.74***(0.62-0.87) | 1.03 (0.98-1.08) | 1.04 (0.99-1.09) | 0.81*(0.67-0.97) | 1.00 (0.95-1.06) | 1.05 (1.00-1.10) | 1.04 (0.87-1.24) | 1.20***(1.14-1.26) | 1.09***(1.04-1.14) |
| OBC | 0.99 (0.84-1.18) | 1.06**(1.02-1.12) | 0.94**(0.90-0.98) | 0.89 (0.73-1.08) | 1.07*(1.01-1.12) | 0.94*(0.90-0.99) | 1.19(0.98-1.43) | 1.12***(1.06-1.18) | 0.86***(0.82-0.90) |
| **Assets index** |  |  |  |  |  |  |  |  |  |
| Poor | Ref. | Ref. | Ref. | Ref. | Ref. | Ref. | Ref. | Ref. | Ref. |
| Middle | 0.84*(0.71-1.00) | 1.02 (0.97-1.07) | 1.01 (0.97-1.06) | 0.86 (0.72-1.04) | 1.02 (0.97-1.08) | 0.99 (0.95-1.04) | 0.75***(0.63-0.89) | 0.96 (0.91-1.01) | 0.94**(0.90-0.98) |
| Rich | 0.72**(0.57-0.92) | 1.05 (0.99-1.12) | 1.01 (0.96-1.07) | 0.70**(0.53-0.92) | 1.07 (1.00-1.14) | 0.96 (0.90-1.02) | 0.69**(0.53-0.89) | 1.00 (0.94-1.07) | 0.87***(0.83-0.92) |
| **Piped drinking water ^d^** |  |  |  |  |  |  |  |  |  |
| No | Ref. | Ref. | Ref. | Ref. | Ref. | Ref. | Ref. | Ref. | Ref. |
| Yes | 0.81**(0.70-0.93) | 0.95**(0.91-0.99) | 0.95**(0.92-0.98) | 0.82**(0.70-0.95) | 1.04 (0.99-1.08) | 1.01 (0.97-1.05) | 0.76***(0.66-0.88) | 1.04 (0.99-1.08) | 1.01 (0.97-1.04) |
| **Toilet facility ^e^** |  |  |  |  |  |  |  |  |  |
| Unimproved | Ref. | Ref. | Ref. | Ref. | Ref. | Ref. | Ref. | Ref. | Ref. |
| Improved | 1.17 (0.97-1.41) | 0.93**(0.88-0.98) | 1.00 (0.95-1.05) | 1.10 (0.90-1.35) | 0.89***(0.84-0.94) | 0.94*(0.89-0.99) | 0.98 (0.81-1.19) | 0.84***(0.80-0.88) | 0.90***(0.85-0.94) |
| **Community open defecation** | 2.07***(1.63-2.63) | 1.46***(1.37-1.56) | 1.41***(1.30-1.53) | 2.11***(1.62-2.74) | 1.44***(1.34-1.55) | 1.33***(1.22-1.45) | 3.12***(2.44-3.98) | 1.76***(1.64-1.88) | 1.61***(1.49-1.75) |
| **Cooking fuel ^f^** |  |  |  |  |  |  |  |  |  |
| Unclean | Ref. | Ref. | Ref. | Ref. | Ref. | Ref. | Ref. | Ref. | Ref. |
| Clean | 1.11 (0.91-1.37) | 0.99 (0.95-1.04) | 0.97 (0.93-1.01) | 1.26* (1.01-1.59) | 1.03 (0.98-1.09) | 1.04 (0.99-1.08) | 1.09 (0.87-1.36) | 1.07*(1.01-1.12) | 0.94**(0.91-0.98) |

_Category 2:_ _mild and non-anaemic dyad; Category 3: severe/moderate and non-anaemic; Category 4: mild and moderate/severely anaemic dyad_

_Level of significance ***p < 0.001, **<p < 0.01, *p < 0.05;_

**^a^** _Primary: schooling for 1-5 years, secondary: schooling for 6-12 years, higher: schooling for >12 years;_

**^b^** _Mass media exposure was created by summing the frequency of exposure to the three media types: reading newspapers/magazines, listening to the radio, and watching TV and then categorizing them as 'no' for zero exposure, 'low' for exposure to one medium, and 'high' for exposure to more than one medium_

**^c^** _Morbidity encompasses at least one of the three conditions: diarrhea, fever and acute respiratory infection (ARI)_

**^d^** _Piped drinking water includes piped water, public taps/standpipes_

**^e^** _Improved toilet facilities include any non-shared toilet of the following types: flush/pour flush toilets to piped sewer systems, septic tanks, pit latrines, or an unknown destination; ventilated improved pit (VIP)/biogas latrines; pit latrines with slabs; and twin pit/composting toilets_

**^f^** _Clean cooking fuel includes electricity, LPG/natural gas, biogas_

**Table S5:** **PAF for the risk factors of anaemia among mother-child dyads (both anaemic) in India**

|  | **NFHS-3** | **NFHS-4** | **NFHS-5** | **Rate of change ^a^** | **Realistic scenario ^b^** | **Optimistic scenarios ^c^** | |
| --- | --- | --- | --- | --- | --- | --- | --- |
|  | 2006 | 2016 | 2021 |  | 2030 | 3% | 5% |
|  | **Panel A: Future prevalence scenario estimates** | | | | | | |
| Higher education (>12 years schooling) | 8.2 | 11.3 | 16.1 | 0.05 | 23.5 | 35.6 | 45.0 |
| Weekly consumption of egg/fish/meat | 47.3 | 50.0 | 54.0 | 0.01 | 58.4 | 71.9 | 79.2 |
| Daily consumption of green vegetables | 64.4 | 46.7 | 53.2 | 0.01 | 57.0 | 58.5 | 59.2 |
| 100+ days IFA consumption | 20.7 | 30.4 | 43.5 | 0.05 | 61.3 | 74.1 | 81.0 |
| ICDS food supplementation | 26.5 | 51.9 | 65.7 | 0.06 | 85.3 | 91.4 | 94.1 |
| Improved toilet facility | 44.5 | 51.4 | 73.8 | 0.03 | 86.7 | 92.3 | 94.7 |
| Community open defecation | 47.9 | 42.5 | 22.0 | -0.05 | 11.4 | 6.0 | 4.1 |
|  | **Panel B: Category 2** | | | | | | |
| Higher education (>12 years schooling) |  |  |  |  | 1.4 | 2.6 | 3.5 |
| Weekly consumption of egg/fish/meat |  |  |  |  | 0.3 | 0.8 | 1.1 |
| Daily consumption of green vegetables |  |  |  |  | 0.2 | 0.3 | 0.4 |
| 100+ days IFA consumption |  |  |  |  | 0.3 | 0.4 | 0.4 |
| ICDS food supplementation |  |  |  |  | 0.2 | 0.3 | 0.3 |
| Improved toilet facility |  |  |  |  | 1.1 | 1.3 | 1.4 |
| Community open defecation |  |  |  |  | 5.2 | 6.2 | 6.5 |
| **All of the above** |  |  |  |  | **8.9** | **12.1** | **13.8** |
|  | **Panel C: Category 3** | | | | | | |
| Higher education (>12 years schooling) |  |  |  |  | 3.0 | 5.4 | 7.2 |
| Weekly consumption of egg/fish/meat |  |  |  |  | 0.7 | 1.9 | 2.6 |
| Daily consumption of green vegetables |  |  |  |  | 0.5 | 0.7 | 0.7 |
| 100+ days IFA consumption |  |  |  |  | 1.3 | 1.9 | 2.2 |
| ICDS food supplementation |  |  |  |  | 0.2 | 0.2 | 0.3 |
| Improved toilet facility |  |  |  |  | 2.2 | 2.6 | 2.7 |
| Community open defecation |  |  |  |  | 6.0 | 7.1 | 7.5 |
| **All of the above** |  |  |  |  | **13.9** | **19.5** | **22.7** |
|  | **Panel D: Category 4** | | | | | | |
| Higher education (>12 years schooling) |  |  |  |  | 4.3 | 7.6 | 10.1 |
| Weekly consumption of egg/fish/meat |  |  |  |  | 1.4 | 3.2 | 4.2 |
| Daily consumption of green vegetables |  |  |  |  | 1.2 | 1.5 | 1.6 |
| 100+ days IFA consumption |  |  |  |  | 1.2 | 1.8 | 2.0 |
| ICDS food supplementation |  |  |  |  | 0.4 | 0.5 | 0.5 |
| Improved toilet facility |  |  |  |  | 3.0 | 3.5 | 3.7 |
| Community open defecation |  |  |  |  | 8.6 | 10.0 | 10.5 |
| **All of the above** |  |  |  |  | **20.8** | **28.5** | **32.9** |

_Category 2:_ _mild and non-anaemic dyad; Category 3: severe/moderate and non-anaemic; Category 4: mild and moderate/severely anaemic dyad_

**^a^** _Average annual rate of change:_ $({\frac{Pt}{Po})}^{1/t}-1$ _where, Pt = NFHS-5, Po = NFHS-3, t = 15 years;_

**^b^** _Realistic scenario projection based on average annual rate of change between NFHS-3 and NFHS-5_

**^c^** _Optimistic 3 and 5 percent refer to deviation from the average annual rate of change used in the realistic scenario_
